# IgG index and not serum 25-hydroxyvitamin D is associated with age at onset of multiple sclerosis

**DOI:** 10.1101/2021.05.31.21258017

**Authors:** Pernilla Stridh, Ingrid Kockum, Jesse Huang

**Author notes:** **Correspondence** Jesse Huang, Centrum for Molecular Medcine, L8:05, SE-171 76 Stockholm, Sweden. Equal contribution (Ingrid Kockum, Jesse Huang).

## Abstract

**Background:** Vitamin D deficiency is associated with an increased risk of multiple sclerosis (MS). However, its effect on the age of disease onset remains unclear. This study examines the relationship between serum 25 hydroxyvitamin D (25(OH)D) levels and age of first symptom onset among recently diagnosed MS patients.

**Method:** Serum 25(OH)D was measured from forty MS patients sampled near disease onset. After correcting seasonal variability, a multivariable linear regression was used to examine associations to age at onset.

**Results:** Serum 25(OH)D was not correlated with age at onset (P>0.5). However, CSF IgG index was lower among patients with later disease onset (β=-5.35, P=0.028). Furthermore, we observed bias resulting from non-random distribution of sampling by season, which after correcting indicates a higher 25(OH)D level among patients sampled at relapse compared to remission, instead of lower as previously reported.

**Conclusion:** In summary, serum 25(OH)D was not associated with the age at onset.

## Introduction

Vitamin D is essential for proper immune regulation, and long-term deficiencies commonly due to inadequate sun exposure and dietary supplementation have been associated with increased autoimmunity [1]. Low sun exposure has also been associated with an increased risk of multiple sclerosis (MS) [2,3], a chronic neuroinflammatory disease of the central nervous system. Geographic gradients in MS prevalence, corresponding with latitude and ultraviolet radiation (UVR) levels, has been consistently reported in previous studies [4-6]. Emphasis on early childhood exposures indicates vitamin D is likely involved in the early stages of disease development [3,7]; however, its direct influence on the age at onset or the incidence of pediatric-onset disease remains unclear [3,8,9]. This study investigates the association between vitamin D and age at first symptom onset among recently diagnosed MS patients while also examining effects from seasonal variability using data from Soilu-Hänninen et al. [10].

## Material and Methods

The study cohort and all sampling and analysis protocols have been previously described [10]. In summary, forty MS patients were enrolled from two Finnish neurology clinics between 2000 and 2003, with serum sampled at or near disease onset. Serum 25 hydroxyvitamin D (25(OH)D, nmol/L), a stable precursor commonly used to assess overall vitamin D deficiency, was determined using a commercial I25 radioimmunoassay. Cerebrospinal fluid (CSF) was sampled to determine the CSF IgG index and the presence of oligoclonal bands (OCB). Summary statistics of the study cohort are provided in Table 1.

**Table 1.** Summary characteristics of MS patients.

| Variable | Mean ± SD [range] / n (%) |
| --- | --- |
| N | 40 |
| + Relapsing-remitting disease | 38 (95%) |
| + Primary progressive disease | 2 (5%) |
| Female | 32 (80%) |
| Age (years) | 36.5 ± 9.0 [18-53] |
| Age at first symptom (years) | 33.8 ± 9.2 [18-53] |
| Age at diagnosis (years) | 36.7 ± 9.1 [18-53] |
| Disease duration (years) | 2.6 ± 3.5 [0-13] |
| Disease duration, ≤ 1 year | 24 (60%) |
| EDSS ≥ 2 | 14 (35%) |
| OCB presence | 32 (82.1%) |
| CSF IgG Index | 1.15 ± 0.76 [0.40-4.82] |
| Serum 25(OH)D level (nmol/L) | 49.6 ± 19.4 [17.0-95.0] |
| Serum 25(OH)D level (adj)* (nmol/L) | 47.8 ± 17.7 [4.4-77.6] |

\* adj. serum 25(OH)D levels adjusted for seasonal variation.

Serum 25(OH)D levels naturally oscillate due to seasonal variation in sun/UV-B exposure, an important source for vitamin D production in addition to diet and supplementation. Levels tend to peak in the summer and drop in the winter. As previously detailed [7], 25(OH)D levels were adjusted for seasonal variability using a periodic regression model of sin(2πx/12) + cos(2πx/12), with “x” defined by the month of sampling. Residuals were then added to the mean determined by the model to calculate the season-adjusted 25(OH)D level.

Associations were analyzed using either a Student t-test or a multivariable linear regression model adjusting for sex, age at sampling, and other relevant disease characteristics. The expanded disability status scale (EDSS), assessed at the time of sampling, was dichotomized by ≥2, corresponding to at least minimal disability in one functional system. All statistical analyses were performed using R v.4.0.3 (Vienna, Austria).

## Results

**Figure 1** shows the periodic seasonal variability in serum 25(OH)D levels, with the highest levels observed in the summer and lowest in late winter.

**Figure 1.**
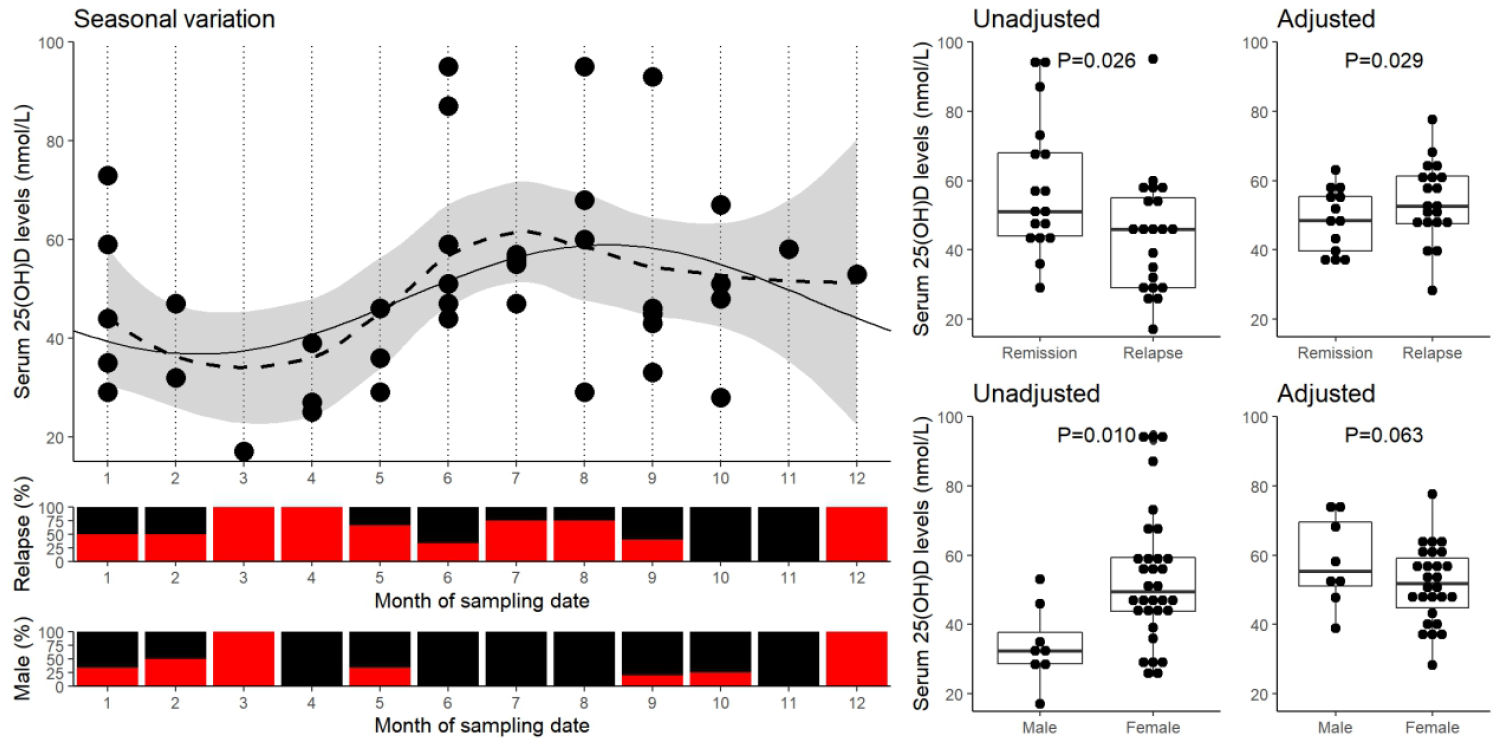
Seasonal variation of serum 25(OH)D levels and its effect on the sampling of relapse/remission patients. The scatter-plot illustrates the seasonal variation in serum 25(OH)D levels among all MS patients, along with the proportion sampled during relapse and of male sex (red) for each month below. A smoothed fitted line (dotted, loess method) with 95% confidence interval (grey interval) are plotted along with fitted periodic regression model of sin(2πx/12) + cos(2πx/12) (solid). Reported significance are of the difference between relapse and remission MS patients determined by a Student t-test for either unadjusted or seasonal variation adjusted serum 25(OH)D levels.

There is a higher proportion of MS patients sampled during relapse between March and May. The risk of sampling a patient undergoing relapse within the period as determined by Fischer’s exact test is OR=6.132 (95% confidence interval=0.63-312, P=0.10). Furthermore, this period corresponds with a naturally lower serum 25(OH)D, which explains the previous observation of lower serum 25(OH)D levels among those sampled during relapse compared to remission (β=-13.9, SE=6.0, P=0.026) [10]. Following correction of seasonal variation (**Figure 1**, right panel), serum 25(OH)D levels among MS patients sampled at relapse were higher than at remission (β=12.0, SE=5.3, P=0.029).

A similar effect was observed when comparing 25(OH)D levels by sex. There was a higher proportion of females between June and August (P=0.03) and a moderately higher proportion of males in the winter between December and March (P=0.059). This also results in a switch in effect (uncorrected: β=19.3, SE=7.1, P=0.01), with females having lower 25(OH)D than males (corrected: β=-13.0, SE=6.8, P=0.06).

Serum 25(OH)D concentration at sampling was not associated with age at onset (P>0.5), even after correcting for sex and other disease characteristics (**Table 2**). Similarly, Pearson and Spearman correlation coefficients were only ρ=0.02 and ρ=0.08, respectively. However, CSF IgG index was negatively correlated with age at onset (P<0.03), and after adjusting for age, sex, and other disease characteristics, IgG index was also positively associated with serum 25(OH)D levels (β=9.2, SE=4.0, P=0.03; **Table 3**).

**Table 2.** Multivariable linear regression of age at MS symptom onset on serum 25(OH)D levels adjusted for season of sampling and disease characteristics.

| Variables | Univariable |  |  | Multivariable* |  |  |
| --- | --- | --- | --- | --- | --- | --- |
| | $\beta$ | SE | P | $\beta$ | SE | P |
| Age | 0.95 | 0.06 | 8.4E-18 | - | - | - |
| Sex, female | 3.67 | 3.65 | 0.32 | 6.39 | 4.23 | 0.14 |
| Serum 25(OH)D - Unadj. | 0.05 | 0.08 | 0.52 | - | - | - |
| Serum 25(OH)D - Adj | 0.01 | 0.08 | 0.91 | 0.06 | 0.10 | 0.56 |
| Relapsing activity | 0.88 | 3.05 | 0.77 | -0.77 | 3.41 | 0.82 |
| OCB (+) | -4.19 | 3.85 | 0.28 | 1.02 | 4.32 | 0.81 |
| CSF IgG index | **-4.79** | **1.80** | **0.011** | **-5.35** | **2.31** | **0.028** |
| EDSS $\geq 2$ | -0.13 | 3.10 | 0.96 | 0.72 | 3.32 | 0.83 |

Associations reaching statistical significance (P<0.05) are highlighted in bold.
\* All the variables listed were included in the final model.

**Table 3.** Multivariable linear regression of serum 25(OH)D levels.

| Variable | Unadjusted |  |  |  |  |  | Season adjusted |  |  |  |  |  |
| --- | --- | --- | --- | --- | --- | --- | --- | --- | --- | --- | --- | --- |
|  | Univariable |  |  | Multivariable* |  |  | Univariable |  |  | Multivariable* |  |  |
| | $\beta$ | SE | P | $\beta$ | SE | P | $\beta$ | SE | P | $\beta$ | SE | P |
| Age | 0.25 | 0.35 | 0.47 | 0.11 | 0.36 | 0.75 | 0.046 | 0.32 | 0.89 | 0.196 | 0.321 | 0.55 |
| Sex, female | **19.28** | **7.12** | **0.010** | 14.41 | 8.64 | 0.11 | -13.00 | 6.78 | 0.063 | -6.36 | 7.75 | 0.42 |
| Relapsing activity | **-13.90** | **5.98** | **0.026** | -11.50 | 6.14 | 0.071 | **12.04** | **5.29** | **0.029** | **12.89** | **5.51** | **0.026** |
| OCB (+) | -1.73 | 7.75 | 0.82 | 8.00 | 8.29 | 0.34 | 2.06 | 7.20 | 0.78 | -7.87 | 7.44 | 0.30 |
| CSF IgG index | -1.66 | 4.11 | 0.69 | -6.07 | 4.45 | 0.18 | 3.46 | 3.72 | 0.36 | **9.24** | **3.99** | **0.028** |
| EDSS $\geq 2$ | -3.15 | 6.50 | 0.63 | 5.37 | 6.39 | 0.41 | 4.09 | 5.92 | 0.49 | -6.01 | 5.74 | 0.30 |

Associations reaching statistical significance ( $P < 0.05$ ) are highlighted in bold. Adjusted serum 25(OH)D levels were adjusted for seasonal variability using a periodic regression model of $\sin(2\pi x/12) + \cos(2\pi x/12)$ , with “x” defined by the month of sampling.
\* All the variables listed were included in the final model.

## Discussion

Our findings show no association between serum 25(OH)D levels and age at MS onset, consistent with previous studies [3,11]. CSF IgG index was associated with age at onset, but this may be due to an association with age, as samples were taken near onset. IgG index have previously been shown to be negatively correlated with age among healthy individuals [12]. However, correlation with age was mainly attributed to a drop in IgG index above 45 years of age, and therefore unlikely to explain the association to age at onset as most patients in this cohort were below 45 years old. Our findings are also supported by an international multi-cohort study observing a similar negative correlation between IgG index and age at onset [13]. However, this association will require further validation in consideration of both age and disease duration.

Relapsing disease and sex were partly dependent on sampling time in this cohort resulting in a bias affecting the previously reported associations with serum vitamin-D levels [10]. Although this could be a random occurrence, annual fluctuations in vitamin D and sun exposure have been associated with relapse rate [14], emphasizing the importance of correcting vitamin D measures for the time of sampling in future studies, particularly with a small sample size. It also showcases the effect of bias that occurs from non-randomized sampling where exploratory variables are confounded by variability in sampling protocol, in this case, resulting in a significant association in the opposite direction.

Although patients were sampled near the first symptom onset, measures may not represent actual disease onset due to a likely period of sub-clinical disease activity. However, a study looking at predicted lifelong residential UVB exposure observed no association to age at MS onset [15]. Genetic association studies and the recent popularity of genetic instruments may provide a reasonable proxy of vitamin D levels prior to onset. Still, a recent Danish study showed no association between age at onset and genetic susceptibilities to vitamin D deficiency, further refuting the role of vitamin D with age at MS onset [8].

In conclusion, these findings indicate serum vitamin D levels were not associated with age at onset, and that it is crucial to correct for seasonal variation of vitamin D to prevent misrepresented conclusions resulting from potential sampling bias.

## Data Availability

The data used in this study are available through https://doi.org/10.1191/1352458505ms1157oa.

## Abbreviations

25(OH)D: 25-hydroxyvitamin D_3_
CSF: cerebrospinal fluid
EDSS: expanded disability status scale
MS: multiple sclerosis
OCB: oligoclonal band
PPMS: primary progressive multiple sclerosis
RRMS: relapsing-remitting multiple sclerosis
UV: ultraviolet

## Acknowledgments

We would like to thank Soilu-Hanninen and collegues for the data used in this study [10]. PS was supported by a grant from Magreta af Ugglas foundation, IK and JH were supported by a Horizon 2020 EU grant (MultipleMS project number 733161). JH was partly supported by an endMS Doctoral Studentship (EGID:3045) from the Multiple Sclerosis Society of Canada.

